# Correlation of SARS-CoV-2 Breakthrough Infections to Time-from-vaccine; Preliminary Study

**DOI:** 10.1101/2021.07.29.21261317

**Authors:** Barak Mizrahi, Roni Lotan, Nir Kalkstein, Asaf Peretz, Galit Perez, Amir Ben-Tov, Gabriel Chodick, Sivan Gazit, Tal Patalon

## Abstract

The short-term effectiveness of a two-dose regimen of the BioNTech/Pfizer mRNA BNT162b2 severe acute respiratory syndrome coronavirus 2 (SARS-CoV-2) vaccine was widely demonstrated. However, long term effectiveness is still unknown. A nationwide vaccination campaign was initiated early in Israel, allowing for a real-world evaluation of the interaction between protection and time-from-vaccine. The Delta (B.1.617.2) variant became the dominant strain in Israel in June 2021, as Israel is currently experiencing a new surge of cases. Leveraging the centralized computerized database of Maccabi Healthcare Services (MHS), we assessed the correlation between time-from-vaccine and incidence of breakthrough infection. We found that the risk for infection was significantly higher for early vaccinees compared to those vaccinated later. This preliminary finding should prompt further investigagions into long-term protection against different strains, and prospective clinical trials to examine the effect of a booster vaccine against breakthrough infection.

## Main

A two-dose regimen of the BioNTech/Pfizer mRNA BNT162b2 severe acute respiratory syndrome coronavirus 2 (SARS-CoV-2) vaccine was demonstrated to be highly effective in preventing infection and symptomatic COVID-19, both in clinical trials^1,2^ and real-world settings^3,4^. However, long term effectiveness is still unknown. Different studies examined the immunological response over time^5–7^, though the correlation between antibody dynamics and clinical protection remains undefined. Additionally, differentiating between time-from-vaccine and vaccine effectiveness across different strains^8–10^ is challenging, as new Variants of Concern (VOC) are rapidly identified.

The Delta (B.1.617.2) variant, initially identified in India and now globally detected, is currently the dominant strain in Israel. In light of the recent surge of cases in Israel, many of which among vaccinated individuals^11^, concerns of reduced vaccine efficacy against the Delta variant have surfaced, including official reports of decreased protection^12^. Contrastingly, other studies report only modest differences in vaccine effectiveness^13^ and substantial antibody response to the Delta variant.^14^

Data regarding the duration of protection are essential for effective resource allocation and vaccine administration, such as the need and urgency of a third dose.^15,16^ Israel’s rapid rollout of the mass vaccination campaign allows us to investigate the correlation between time-from-vaccine and vaccine effectiveness against the Delta variant.

To this end, we conducted a retrospective cohort study comparing the incidence rates of breakthrough infections between early and late vaccinees, using data from Maccabi Healthcare Services (MHS), Israel’s second largest Health Maintenance Organization, which covers 2.5 million members (25% of the population) and provides a representative sample of the Israeli population.

The study population consisted of all MHS members aged 16 and above who received the second dose of the vaccine between January and April 2021. Individuals were considered fully vaccinated if they received two doses of the BNT162b2, the second one administered within the 21-to-28-day interval set by national guidelines. The minority who did not follow the guidelines included those infected after the first dose or those suffering an intercurrent illness that delayed the administration of the second dose.

Individuals were excluded from the study if they had a positive SARS-CoV-2 polymerase chain reaction (PCR) assay test result prior to the start of the study period or disengaged from MHS for any reason between January and April.

Individual level data of the study population included age, sex, city of residence, last documented body mass index (BMI) (categorized as normal weight <25, overweight 25-30 and obese >30), and socioeconomic status (SES), on a scale from 1 (lowest) to 10. SES index is based on several parameters including household income, educational qualifications, household crowding, material conditions, and car ownership. Data collected also included sinformation of chronic diseases from MHS’ automated registries, including cardiovascular diseases^17^,hypertension, diabetes^18^, chronic kidney disease (CKD)^19^, chronic obstructive pulmonary disease (COPD), inflammatory bowel disease (IBD) and immunocompromised conditions, as well as data on cancer from the National Cancer Registry^20^. Additionally, dates of the first and second dose of the vaccine (if received) and results of any polymerase chain reaction (PCR) tests for SARS-CoV-2, all recorded centrally in MHS, were included in the analysis.

To assess the correlation between time-from-vaccine and afforded protection against breakthrough infection, two logistic regression models were applied. In both models, the outcome was defined as a positive SARS-CoV-2 PCR test recorded between June 1^st^ and July 27^th^, the date of analysis.

In the first model, we addressed the time-from-vaccine by grouping individuals into two separate groups of comparison: Early Vaccinees and Late Vaccinees. We defined Early Vaccinees as individuals who received the second dose of the vaccine between January and February 2021 and Late Vaccinees as individuals who received the second dose between March and April 2021. As the mass vaccination campaign first targeted high-risk individuals (e.g., healthcare personnel and persons with comorbidities) and those over the age of 60, we matched each Early Vaccinee to a Late Vaccinee individual in a 1:1 ratio, based on age group (18-39, 40-59 and 60 and over), sex, city of residence and socioeconimoc status. Results were then adjusted for underlying comorbidities, including obesity, cardiovascular diseases, diabetes, hypertension, chronic kidney disease, cancer, COPD, IBD and immunosuppression conditions. We applied logistic regression to calculate the odds ratio (OR) of SARS-CoV-2 infections between the groups with associated 95% confidence intervals (CIs).

In the second model, we addressed the time-from-vaccine by analyzing six distinct groups, comparing individuals according to the month in which they were first considered to be fully vaccinated (the groups were January-February, January-March, January-April, February-March, February-April and March-April). Thereby, we compared the incidence of SARS-CoV-2 breakthrough infection between individuals who were fully vaccinated in January 2021 and those who were fully vaccinated in February 2021 and so on. The same matching and adjustment methods were performed in both models.

Analyses were performed using Python version 3.1 with the stats model package.

Of 1,395,134 MHS members over the age of 16 who received the second dose of the vaccine between January and April of 2021, 1,352,444 were eligible for the study. 27,143 individuals did not receive the second dose according to the guidelines, and 15,547 individuals were tested positive to SARS-CoV-2 prior to the study period.

The incidence rates per 10,000 individuals who received their second dose during the months of January, February, Mar and April were 36.5, 33.65, 23.06 and 16.98, respectively (Figure 1).

**Figure 1.**
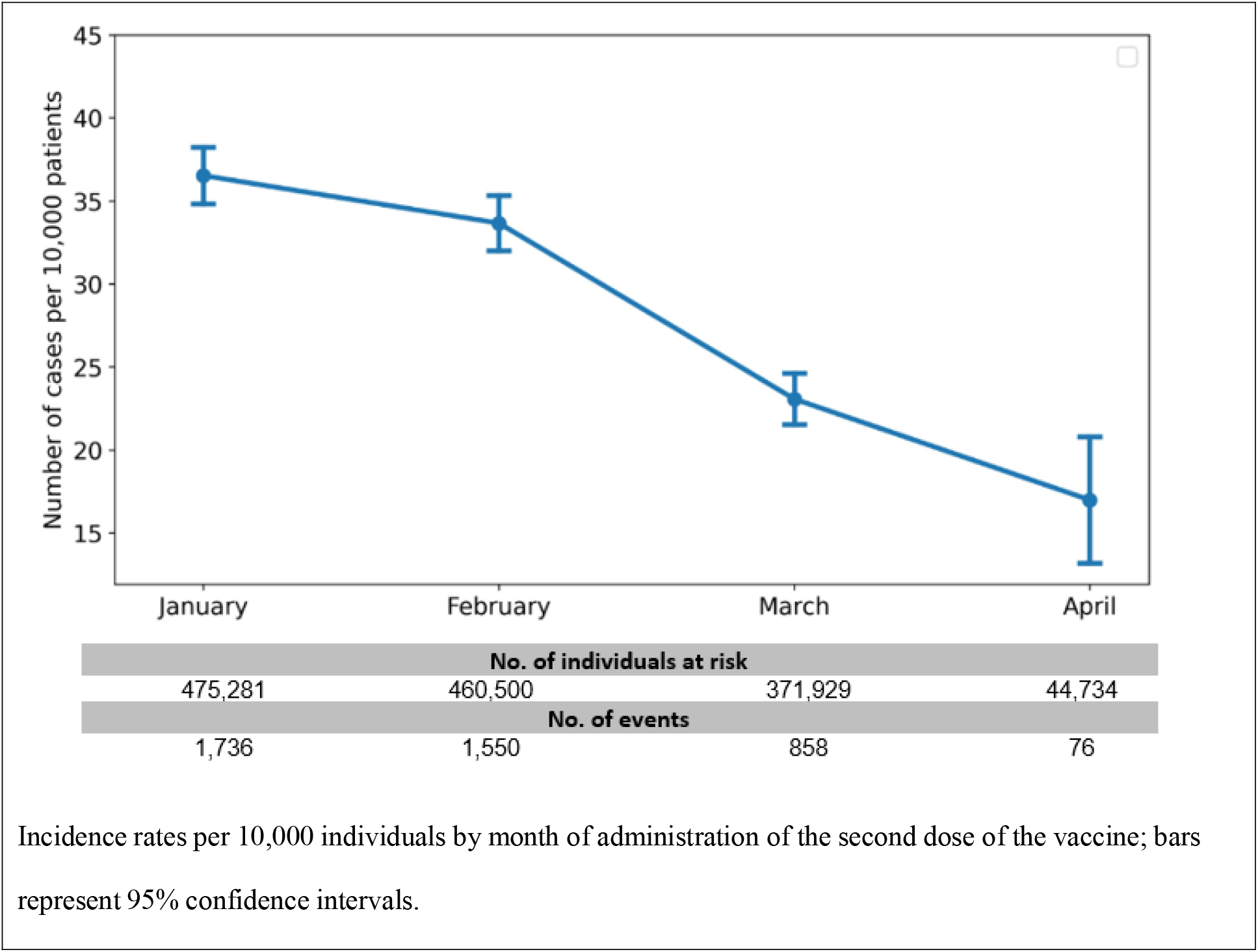
Incidence Rate by Month

In model 1, we matched 329,177 persons in each group. During the follow-up period, 1,911 cases of breakthrough infection were recorded, 1151 of them in the Early Vaccinees group and 760 in the Late Vaccinees group. After adjusting for comorbidities, we found a statistically significant 53% (CI 40–68%) increased risk for breakthrough infection in early Vaccinees (*P*<0.001) (Table 1, Model 1). When stratifying the results by age, we found a similar trend across all age groups.

**Table 1.**
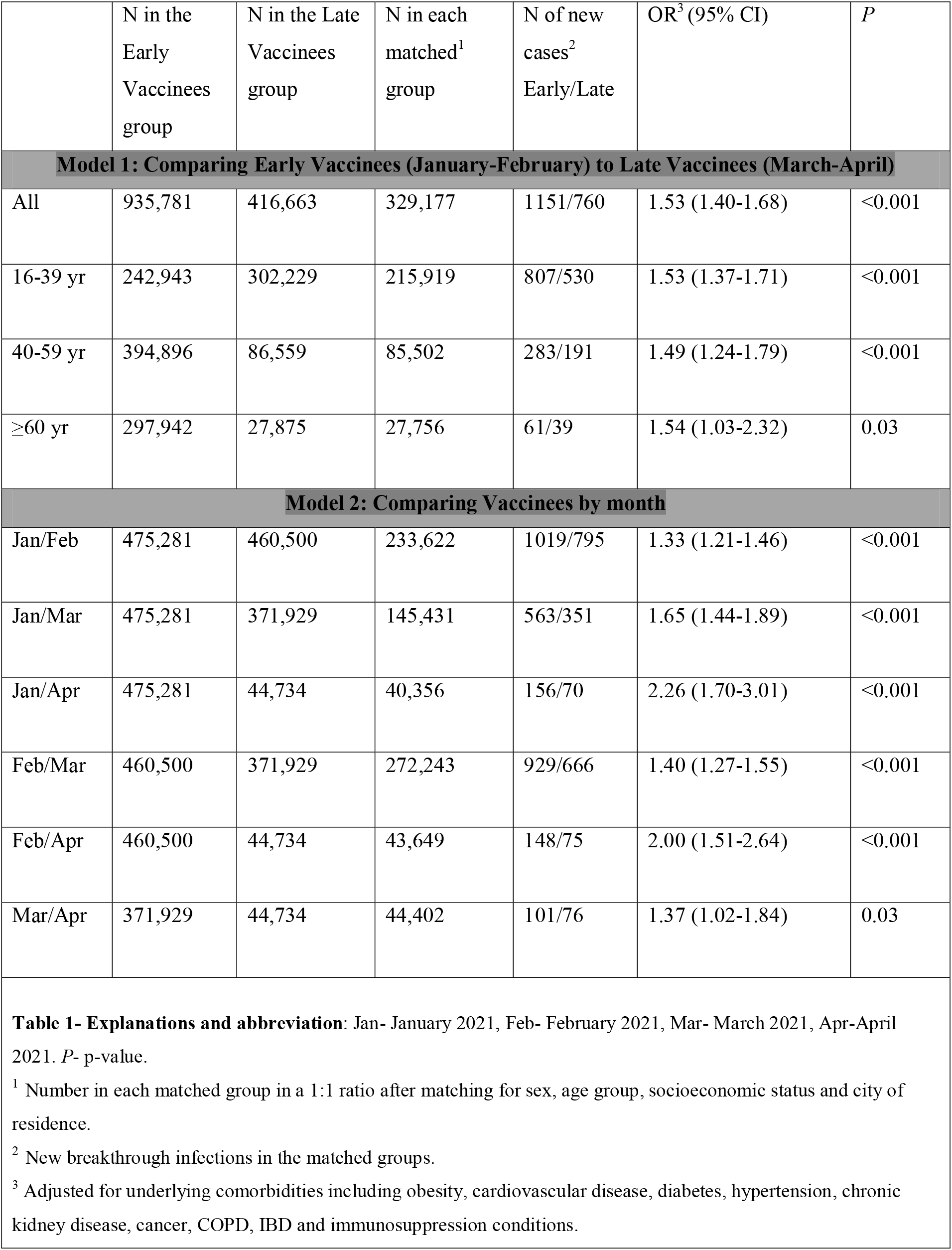
Odds Ratio (ORs) of SARS-CoV-2 Breakthrough Infections Between Early and Late Vaccinee Groups

Explanations and abbreviation: Jan- January 2021, Feb- February 2021, Mar- March 2021, Apr-April 2021. P- p-value.
|  | N in the Early Vaccinees group | N in the Late Vaccinees group | N in each matched<sup>1</sup> group | N of new cases<sup>2</sup> Early/Late | OR<sup>3</sup> (95% CI) | P |
| --- | --- | --- | --- | --- | --- | --- |
| **Model 1: Comparing Early Vaccinees (January-February) to Late Vaccinees (March-April)** |  |  |  |  |  |  |
| All | 935,781 | 416,663 | 329,177 | 1151/760 | 1.53 (1.40-1.68) | <0.001 |
| 16-39 yr | 242,943 | 302,229 | 215,919 | 807/530 | 1.53 (1.37-1.71) | <0.001 |
| 40-59 yr | 394,896 | 86,559 | 85,502 | 283/191 | 1.49 (1.24-1.79) | <0.001 |
| ≥60 yr | 297,942 | 27,875 | 27,756 | 61/39 | 1.54 (1.03-2.32) | 0.03 |
| **Model 2: Comparing Vaccinees by month** |  |  |  |  |  |  |
| Jan/Feb | 475,281 | 460,500 | 233,622 | 1019/795 | 1.33 (1.21-1.46) | <0.001 |
| Jan/Mar | 475,281 | 371,929 | 145,431 | 563/351 | 1.65 (1.44-1.89) | <0.001 |
| Jan/Apr | 475,281 | 44,734 | 40,356 | 156/70 | 2.26 (1.70-3.01) | <0.001 |
| Feb/Mar | 460,500 | 371,929 | 272,243 | 929/666 | 1.40 (1.27-1.55) | <0.001 |
| Feb/Apr | 460,500 | 44,734 | 43,649 | 148/75 | 2.00 (1.51-2.64) | <0.001 |
| Mar/Apr | 371,929 | 44,734 | 44,402 | 101/76 | 1.37 (1.02-1.84) | 0.03 |

<sup>1</sup> Number in each matched group in a 1:1 ratio after matching for sex, age group, socioeconomic status and city of residence.
<sup>2</sup> New breakthrough infections in the matched groups.
<sup>3</sup> Adjusted for underlying comorbidities including obesity, cardiovascular disease, diabetes, hypertension, chronic kidney disease, cancer, COPD, IBD and immunosuppression conditions.

Correspondingly, Model 2 demonstrated higher risks for breakthrough infections in persons who were vaccinated early compared to late in each month-group (Table 1, Model 2). Individuals who were vaccinated in January 2021 had a 2.26-fold increased risk (CI 1.80-3.01) for breakthrough infection compared to individuals who were vaccinated in April 2021 (Figure 1).

In this cohort of MHS members, all of whom are vaccinated with the BioNTech/Pfizer mRNA BNT162b2 vaccine in a two-dose regimen, we identified a significant correlation between time-from-vaccine and afforded protection against SARS-CoV-2 infection. The risk for breakthrough infection was significantly higher for early vaccinees compared to those vaccinated later.

Our study has several important limitations. First, as the Delta variant was the dominant strain in Israel during the study period, the observed decrease in long-term protection afforded by the vaccine against other strains cannot be inferred. Second, we did not measure the effect of vaccination time on symptomatic infection, severe disease or hospitalization. Lastly, the results might be affected by differences between the groups in terms of health behaviors (such as social distancing and mask-wearing), a possible confounder that was not assessed. As chronically-ill patients were given priority for vaccination, confounding by indication should be considered when interpreting the study results; nonetheless, adjusting for obesity, cardiovascular disease, diabetes, hypertension, chronic kidney disease, cancer, COPD, IBD and immunosuppression had only a small impact on the estimate of effect as compared to the unadjusted OR. Therefore, residual confounding by unmeasured facotrs is unlikely.

Taken together, the study suggests a possible relative decrease in the long-term protection of the BNT162b2 vaccine against the Delta variant of SARS-CoV-2. This preliminary finding should be evaluated in future studies, including a comparison to long-term protection against different strains, and prospective clinical trials to examine the effect of a booster vaccine against breakthrough infection.

## Data Availability

Data not available due to ethical restrictions

## Ethics declaration

This study was approved by the MHS Institutional Review Board. Due to the retrospective design of the study, informed consent was waived by the IRB and all identifying details of the participants were removed before computational analysis.

## CRediT Authorship Contributions

Conceptualization: RL, SG, TP

Data curation: RL

Formal Analysis: BM, NK

Methodology: RL, GC

Investigation: BM, NK, RL

Visualization: BM

Project administration: SG, GP, TP

Supervision: SG, TP

Writing – original draft: RL, BM, SG, TP

Writing – review & editing: GC, ABT, AP, GP, TP, GC

